# SARS-CoV-2 Detection in Different Respiratory Sites: A Systematic Review and Meta-Analysis

**DOI:** 10.1101/2020.05.14.20102038

**Authors:** Abbas Mohammadi, Elmira Esmaeilzadeh, Yijia Li, Ronald J. Bosch, Jonathan Z. Li

## Abstract

**Background:** The accurate detection of SARS-CoV-2 through respiratory sampling is critical for the prevention of further transmission and the timely initiation of treatment for COVID-19. There is a diverse range of SARS-CoV-2 detection rates in reported studies, with uncertainty as to the optimal sampling strategy for COVID-19 diagnosis and monitoring.

**Methods:** We performed a systematic review and meta-analysis of studies comparing respiratory sampling strategies for the detection of SARS-CoV-2 RNA. The inclusion criteria were studies that assessed at least two respiratory sampling sites (oropharyngeal swab, nasopharyngeal swab, and sputum) in participants with COVID-19. The percentage positive tests were compared between sampling modalities by constructing a Z-test assuming independence and using the standard errors obtained from the random effects meta-analysis.

**Findings:** From 1039 total studies, we identified 11 studies that met our inclusion criteria, with SARS-CoV-2 testing results from a total of 3442 respiratory tract specimens. Compared to nasopharyngeal swab sampling, sputum testing resulted in significantly higher rates of SARS-CoV-2 RNA detection while oropharyngeal swab testing had lower rates of viral RNA detection. Earlier sampling after symptom onset was associated with improved detection rates, but the differences in SARS-CoV-2 RNA detection by sampling method was consistent regardless of the duration of symptoms.

**Interpretation:** The results support sputum sampling as a primary method of COVID-19 diagnosis and monitoring, and highlight the importance of early testing after symptom onset to increase the rates of COVID-19 diagnosis.

**Funding:** This study was funded in part by the NIH grants U01AI106701 and by the Harvard University for AIDS Research (NIAID 5P30AI060354).

## Introduction

Worldwide, there are now over 3 million confirmed cases and 250,000 deaths due to Coronavirus disease 2019 (COVID-19), caused by the SARS-CoV-2 virus. The most common route of viral transmission is through exposure to respiratory secretions of close contacts^1^ as the respiratory tract represents the major area of viral shedding.^2^ The accurate diagnosis of COVID-19 infection through respiratory sampling is critical for the prevention of further transmission, clinical trial inclusion criteria and the timely initiation of treatment. In addition to its importance in diagnosis, respiratory sampling plays a central role in determining the duration of viral shedding, with implications for clinical management of potentially infectious patients, decisions on the duration of social isolation, and our understanding of viral transmission and pathogenesis.^3^

Nasopharyngeal swabs are one of the most commonly used methods of respiratory secretion sampling for the detection of SARS-CoV-2 viral RNA. However, the use of nasopharyngeal swabs have a number of drawbacks, including that high-quality swab samples are technically challenging to obtain,^4^ nasopharyngeal swabbing increases the risk to healthcare providers due to the frequent induction of reflex sneezing/coughing, and the disruption in the supply of swabs, transport media, and personal protective equipment (PPE). For all of these reasons, there is intense interest in the comparison of nasopharyngeal swab and alternative sampling methods for the detection of SARS-CoV-2 RNA at respiratory sites. In Asia and other parts of the world, oropharyngeal swabs are the preferred method of COVID-19 diagnosis^2,3^ and there is also interest in the study of sputum as an effective, and less invasive method of COVID-19 diagnosis.

In the published literature, there has been a wide variance in the reported SARS-CoV-2 detection rates with each of the diagnostic methods. In COVID-19 diagnosed individuals, the reported SARS-CoV-2 detection rate has ranged from 25% to >70% of collected nasopharyngeal swabs,^5,6^ 32% to 65% for oropharyngeal swabs,^2,5^ and 48% to >90% for sputum.^7,8^ This has led to significant uncertainty and confusion in the field as to the reason behind the disparate testing results and the optimal diagnostic sampling strategy for diagnosis and monitoring of COVID-19 patients. In this study, we performed a systematic review of the literature and meta-analysis to compare the ability of nasopharyngeal swabs, oropharyngeal swabs, and sputum to detect SARS-CoV-2 RNA. The results demonstrate that sputum had the highest rate of SARS-CoV-2 detection and oropharyngeal swab had the lowest. In addition, much of the variability between reported results in the literature can be attributed to differences in the underlying participant population, with the higher levels of detection for those earliest in the COVID-19 disease course.

## Methods

### Search strategy and selection criteria

In this systematic review and meta-analysis, we used the Preferred Reporting Items for Systematic Reviews and Meta-Analyses (PRISMA) to review literatures and report our results (Figure 1). A computerized search was implemented in PubMed, MedRxiv and BioRxiv using a search term, “((COVID OR COVID-19 OR SARS) AND (throat OR nasal OR nasopharyngeal OR oropharyngeal OR oral OR saliva OR sputum OR PCR))”. The search was completed through April 30, 2020. In addition, we reviewed the reference sections of relevant articles. We included studies that assessed at least two respiratory sites of sampling performed by healthcare workers for individuals with confirmed COVID-19. When numerical results were not reported or further clarification was needed, we contacted the corresponding authors for additional information.

**Figure 1.**
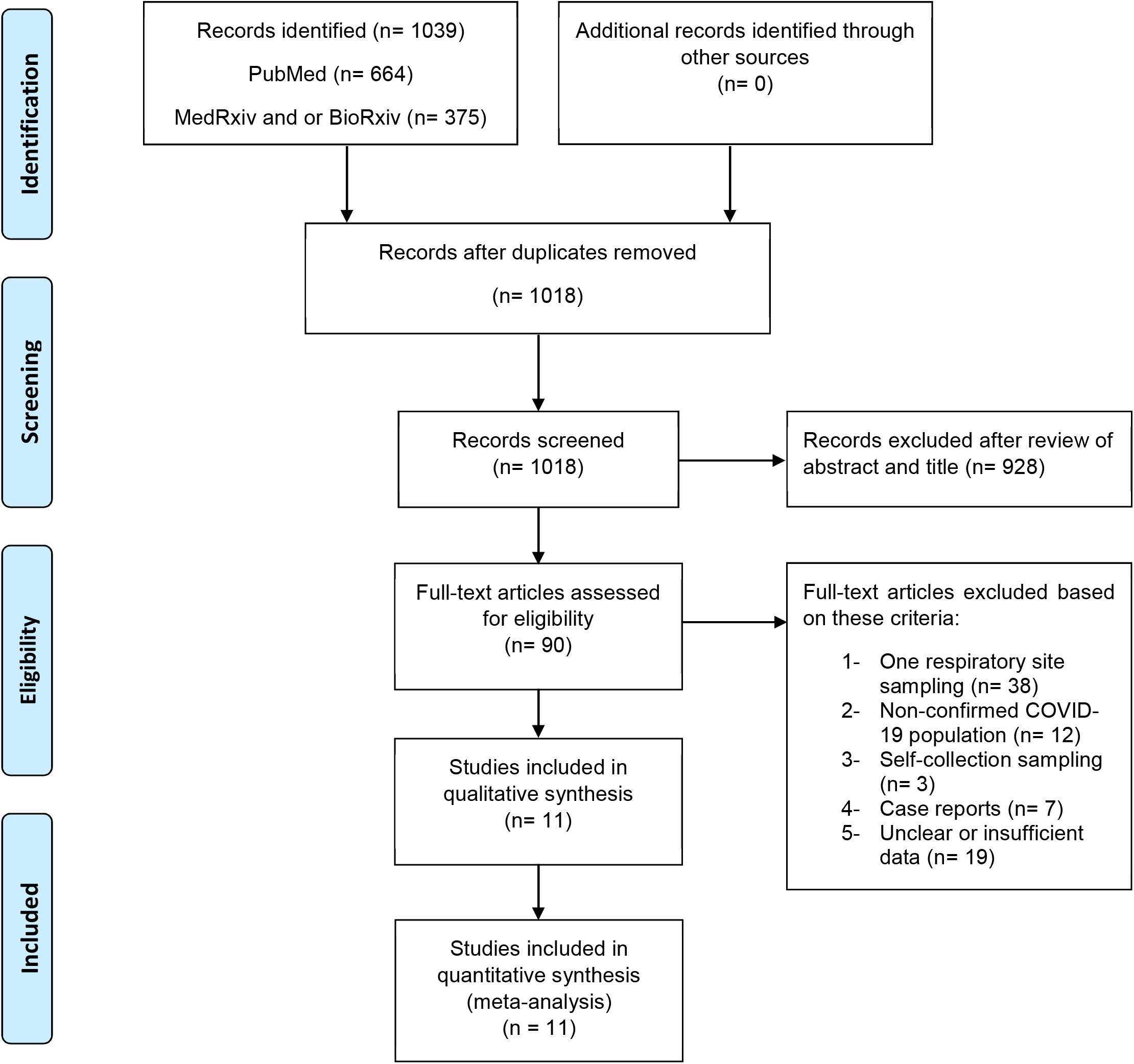
Study profile.

Two authors (AM, EE) screened the citations and three authors (AM, EE, YL) independently extracted data from the included studies. In the 11 studies included in this meta-analysis, we extracted summary estimates from tables or texts from four studies. For the other nine studies, we were able to obtain individual-level data from the manuscripts or study authors.

### Data analysis

The estimated percentages of positive SARS-CoV-2 qPCR tests and the 95% confidence intervals were calculated for each sampling strategy using random-effects meta-analyses for binomial data using Stata Metaprop package.^9^ We also performed an analysis stratified by the duration of symptoms prior to the testing, with results categorized as 0–7 days, 8–14 days, >14 days from symptom onset. We compared the proportion of positive tests between sampling sites by constructing a Z-test assuming independence and using the standard errors obtained from the random effects meta-analysis. For analyses involving studies with small sample size and sensitivity value extremely high (towards 1) or low (towards 0), we incorporated Freeman-Tukey Double Arcsine Transformation method to stabilize the variances for the by-study confidence intervals.

We used inconsistency index (I^2^) test to assess the heterogeneity between each study. Meta-analyses were performed using Stata 13.1 (StataCorp). GraphPad 8.5 (Prism) was used to demonstrate sensitivities at different time points from different sites of respiratory tracts. No adjustments for multiple comparisons were performed.

## Results

From the 1039 studies identified in our search, we excluded 21 duplicates and after screening the abstracts of the remaining articles, 90 full-text articles were obtained for further review (Figure 1). Based on our selection criteria, 79 of those studies were excluded and 11 studies met our inclusion criteria.^1,2,5–7,10–15^ In total, 757 COVID-19 confirmed patients with 3442 respiratory samples were included in this analysis, including results from 1083 oropharyngeal swabs, 1299 nasopharyngeal swabs and 1060 sputum samples. By sampling method, the estimated percentage of positive samples was 43% (95% confidence interval [95% CI]: 34–52%) for oropharyngeal swabs, 54% (95% CI: 41–67%) for nasopharyngeal swabs and 71% (95% CI: 61–80%) for sputum (Figure 2A-C). The rate of SARS-CoV-2 detection was significantly higher in sputum than either oropharyngeal swabs or nasopharyngeal swabs (Figure 2D).

**Figure 2.**
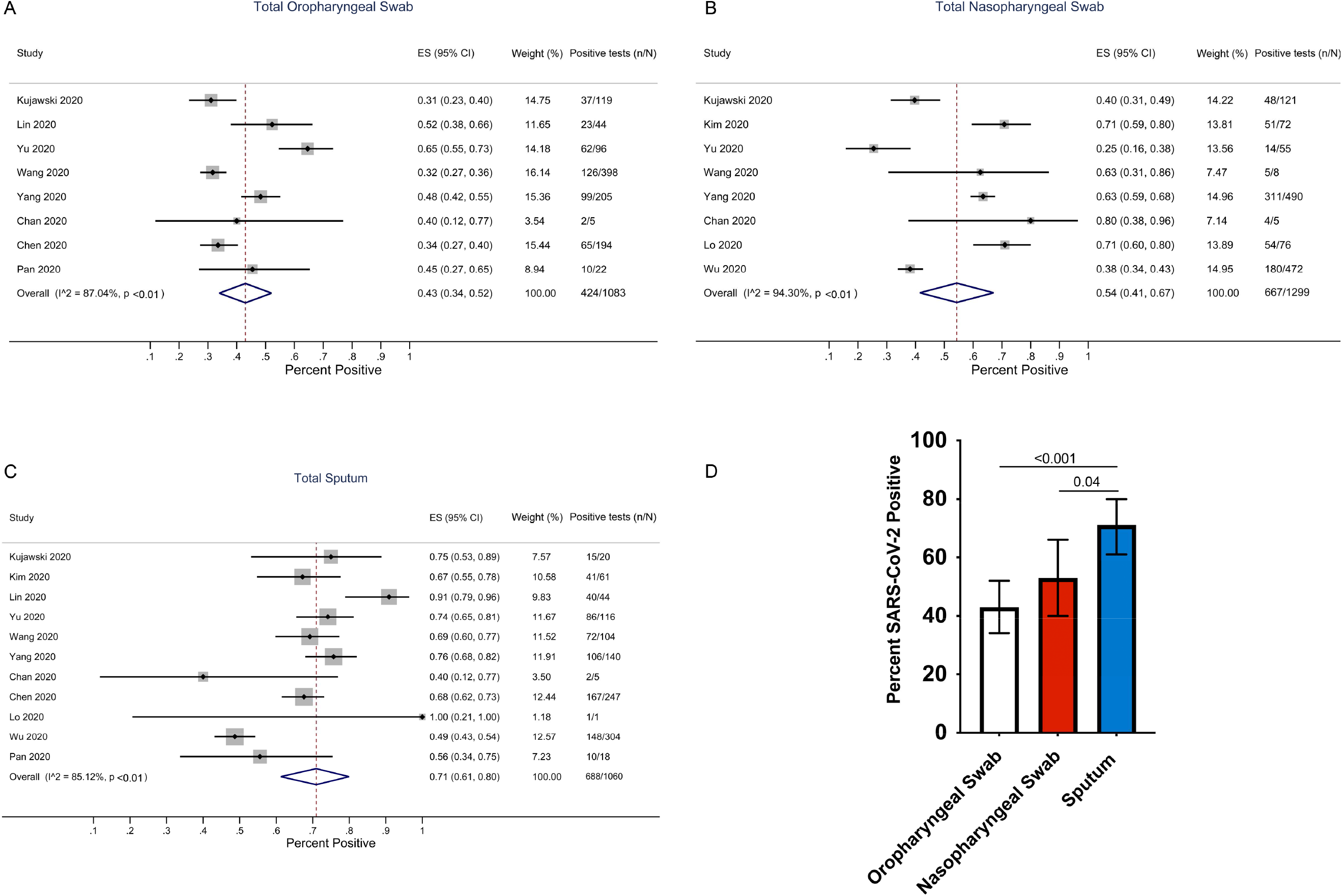
Rates of SARS-CoV-2 detection by three methods of sampling. Forest plots of detection rates for oropharyngeal swabs (A), nasopharyngeal swabs (B), and sputum (C) and in a pooled analysis (D). The error bars in figure 2 D are 95% Confidence Interval (95% CI)

Among the 11 studies, 6 provided details on the time of sampling after symptom onset, including results from 540 oropharyngeal swabs, 759 nasopharyngeal swabs, and 487 sputum samples. For all sampling methods, rates of SARS-CoV-2 detection was highest early after symptom onset. For oropharyngeal swab sampling, the estimated percentage of positive tests were 75% (95% CI: 60–88%) between days 0–7, 35% (95% CI: 27–43%) between days 8–14 and 12% (95% CI: 2–25%) after 14 days from symptom onset (Figure 3A). For nasopharyngeal swabs, the estimated percentage positive was 80% (95% CI: 66–91%), 59% (95% CI: 53–64%) and 36% (95% CI: 18–57%) at 0–7 days, 8–14 days and >14 days after symptom onset, respectively (Figure 3B). For sputum, the estimated percentage positive was 98% (95% CI: 89–100%), 69% (95% CI: 57–80%), and 46% (95% CI: 23–70%) at 0–7 days, 8–14 days, and >14 days after symptom onset, respectively (Figure 3C). For every time period, sputum had the highest percentage of positive results while oropharyngeal swabs had the lowest (Figure 3D).

**Figure 3.**
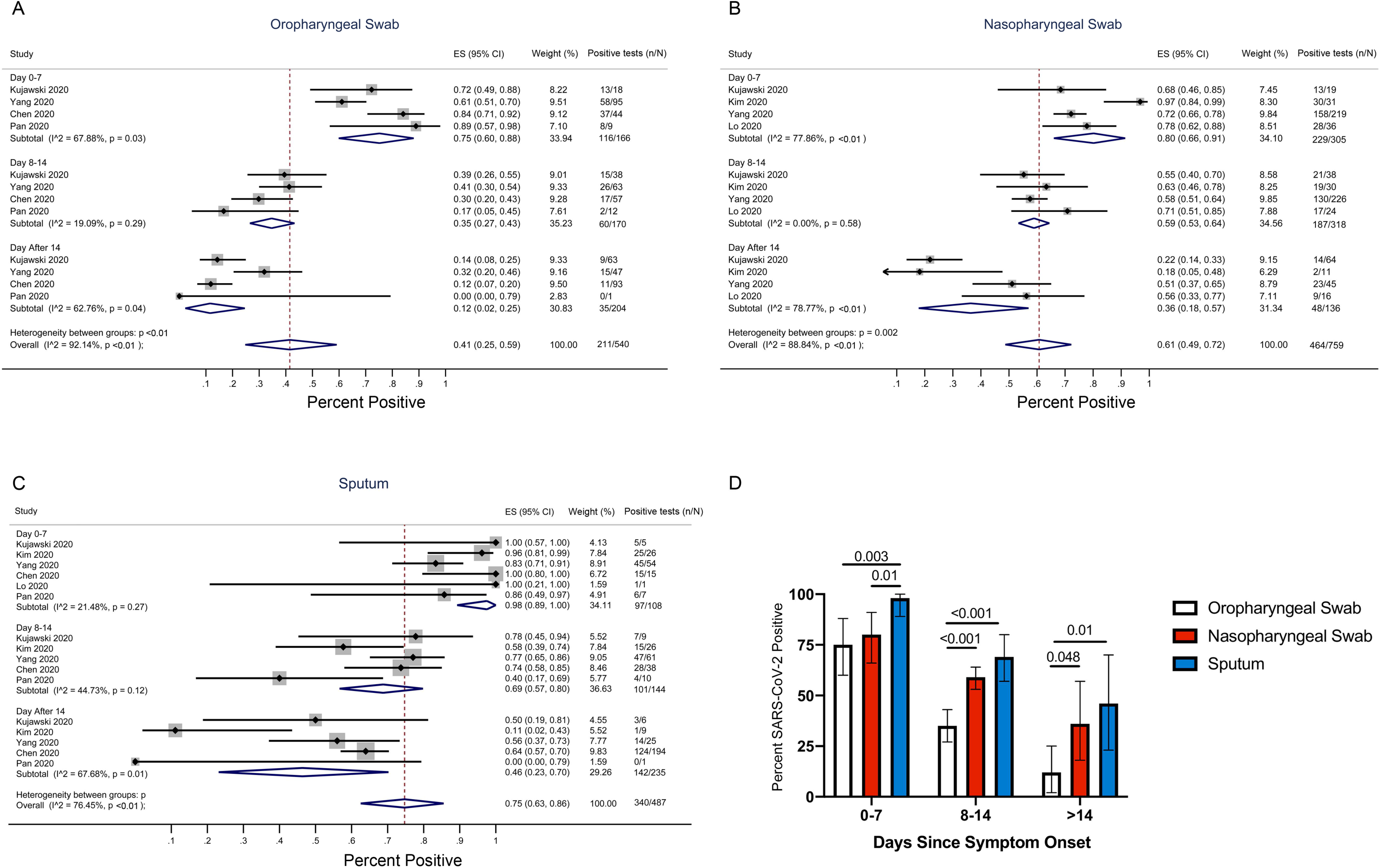
Rates of SARS-CoV-2 detection by three methods of sampling and time since symptom onset. Forest plots of detection rates for oropharyngeal swabs (A), nasopharyngeal swabs (B), and sputum (C) are categorized by days since symptom onset (0–7, 8–14, >14 days) and in a pooled analysis (D). The error bars in figure 2 D are 95% Confidence Interval (95% CI)

**Table 1.** Studies included in the Meta-Analysis.

|  | Country | Age Median<br>(Range) | Sex (M %) | Patients (N) | Total samples |
| --- | --- | --- | --- | --- | --- |
| Kujawski et al (2020)<sup>10</sup> | US | 53 (2-68) | 68 | 12 | 219 |
| Kim et al (2020)<sup>6</sup> | Korea | 40 (20-73) | 54 | 28 | 133 |
| Lin et al (2020)<sup>11</sup> | China | 57 (38-84) | 52 | 52 | 88 |
| Yu et al (2020)<sup>5</sup> | China | 40 (32-63) | 50 | 76 | 267 |
| Wang et al (2020)<sup>2</sup> | China | 44 (5-67) | 68 | 205 | 525 |
| Yang et al (2020)<sup>12</sup> | China | 52 (2-86) | 60 | 213 | 864 |
| Chan et al (2020)<sup>1</sup> | China | 63 (10-66) | 50 | 5 | 15 |
| Chen et al (2020)<sup>13</sup> | China | 36 (2-65) | 64 | 22 | 440 |
| Lo et al (2020)<sup>14</sup> | China | 54 (27-64) | 30 | 10 | 85 |
| Wu et al (2020)<sup>7</sup> | China | 67 ± 9<sup>*</sup> | 55 | 132 | 776 |
| Pan et al (2020)<sup>15</sup> | China | .. | .. | 2 | 40 |

\* Mean age ± SD

In the overall pooled analysis, we detected significant heterogeneity between studies (P<0.001 for all detection methods, Figure 2A-C). Much of the heterogeneity between studies could be accounted for by differences in the participant populations, specifically the timing of symptom onset. In the analysis stratified by days since symptom onset, we observed substantially lower rates of heterogeneity between studies (Figure 3A-C).

## Discussion

In this systematic review and meta-analysis, we combined data from 11 papers that in total reported SARS-CoV-2 RNA testing from a total of 1299 nasopharyngeal swabs, 1083 oropharyngeal swabs, and 1060 sputum samples. The results demonstrate that the rate of sample positivity was highest in sputum specimens and lowest in oropharyngeal swabs. We detected heterogeneity between the reported results by study, much of which could be accounted for by differences in the timing of respiratory sampling. For all three sampling modalities, we found that the likelihood of a positive result declined with longer time since symptom onset. Regardless of the time frame studied, sputum consistently had the highest positive rates while oropharyngeal swabs had the lowest.

Like SARS-CoV, the disease manifestation of SARS-CoV-2 is largely in the lower respiratory tract.^16,17^ Prior studies have noted a higher rate of SARS-CoV-2 detection and viral loads in lower respiratory tract samples, such as bronchoalveolar lavage fluid and endotracheal aspirates, compared to upper respiratory tract specimens.^2,3^ These results are concordant with the far higher density of the SARS-CoV-2 viral target, the human angiotensin-converting enzyme 2 (ACE2) receptor, in pneumocytes and in lower airway epithelial cells compared to epithelial cells in the upper airway.^18,19^ The higher SARS-CoV-2 detection rates in sputum samples likely relate to their origin, which is derived at least partially from the lower respiratory tract. This suggests that sampling of only upper respiratory tract samples, e.g., using nasopharyngeal swabs, may lead to missed diagnosis of COVID-19 and an inaccurate assumption of SARS-CoV-2 viral clearance that dictates the duration of social isolation and return to work policies.

Nasopharyngeal swabs remain the gold-standard for COVID-19 diagnosis in many parts of the world, but their use has a number of drawbacks. First, nasopharyngeal swabs can be technically challenging to obtain^4^ and poor sample quality remains a problem. Second, nasopharyngeal swabbing causes discomfort and frequent reflex sneezing or coughing, and thus requires high-level personal protective equipment for healthcare workers, which are in short supply. Finally, there is a global shortage of nasopharyngeal swabs and transport medium that necessitates the search for potential alternative methods for the sampling of respiratory secretions. In addition to our finding of improved detection of SARS-CoV-2 RNA, the use of sputum sampling removes the need for difficult to obtain swabs, improves patient acceptance, and increases the chances of sample self-collection by the patient, which would decrease the risk to healthcare workers. Furthermore, the higher sensitivity of sputum for the detection of COVID-19 is also supported by previously reported studies on the detection of non-COVID-19 respiratory viruses.^20–22^

One limitation of this meta-analysis is that the majority of participants in these studies enrolled hospitalized patients and it is unclear how the results may differ for individuals with asymptomatic infection or only mild symptoms. However, there are reports that rates of nasopharyngeal, oropharyngeal, and sputum positivity may not be substantially altered in those with mild symptoms versus those with severe disease.^12^ A potential limitation with the use of sputum is that not all COVID-19 patients are able to expectorate sputum, which may be reflected by the lower number of tested sputum samples in the included studies compared to nasopharyngeal or oropharyngeal swabs. Thus, for those who are unable to produce sputum, nasopharyngeal testing may continue to play an important role in the diagnosis or monitoring of COVID-19 patients.

A recent report also suggested that saliva samples had comparable diagnostic accuracy compared to nasopharyngeal swabs, although this will need to be confirmed with additional studies.^23^ Finally, there were differences between studies in the qPCR assay and patient selection (Supplemental Tables 1 and 2) that likely accounted for some of the heterogeneity, the results were generally consistent within most studies and we were able to dramatically reduce the detected heterogeneity of the studies by comparing results by timing of symptom onset.

In summary, this systematic review and meta-analysis demonstrates that compared to nasopharyngeal swab sampling, sputum testing resulted in significantly higher rates of SARS-CoV-2 RNA detection while oropharyngeal swab testing had lower rates of viral RNA detection. Earlier sampling after symptom onset was associated with improved detection rates, but the differences in SARS-CoV-2 RNA detection was consistent between sampling strategies regardless of the duration of symptoms. The results support the use of sputum testing as a primary method of COVID-19 diagnosis and monitoring.

## Data Availability

Our data will be available upon request.

## Contributors

AM, EE, YL, and JL performed the literature review and data abstraction. AM, EE, YL, RB, and JL were involved in the statistical analysis. All authors contributed to the writing and editing of the manuscript.

## Acknowledgements

We would like to thank Dr. Myoung-don Oh for providing additional data from his manuscript and the authors of contributing studies and MedRxiv for making the findings available through publicly accessible pre-prints.

